# Factors associated with increased mortality in critically ill COVID-19 patients in a Mexican public hospital: the other faces of health system oversaturation

**DOI:** 10.1101/2021.03.04.21252084

**Authors:** Mariana Jocelyn Macías Guzmán, Alejandro Castillo-Gonzalez, Jose Lenin Beltran Gonzalez, Mario González Gámez, Emanuel Antonio Mendoza Enciso, Itzel Ovalle Robles, Andrea Lucia García Díaz, César Mauricio Gutiérrez Peña, Lucila Martinez Medina, Victor Antonio Monroy Colin, Arreola Guerra Jose Manuel

**Author notes:** **Corresponding Authors:** José Manuel Arreola Guerra, MD, M. Sc., Ph.D, Head, Department of Investigation, Nephrologist, and Associate Professor of Internal Medicine., Hospital Centenario Miguel Hidalgo, Av Gómez Morín y Av La Estación, La Alameda, Aguascalientes, guascalientes Mexico., C.P. 20259, **Mario Gonzalez Gamez MD**, Department of Internal Medicine and Infectologist, Hospital Centenario Miguel Hidalgo, Av Gómez Morín y Av La Estación, La Alameda, Aguascalientes, Aguascalientes Mexico., C.P. 20259.

## Abstract

**BACKGROUND:** The lethality rate of COVID-19 in Mexico is one of the highest worldwide, but in-hospital factors associated with this increased rate have yet to be explored. This study aims to evaluate those factors that could be associated with mortality at 28-days in critically ill COVID-19 patients in Mexico.

**METHODS:** This is a retrospective analysis of the patients included in the clinical trial (NCT04381858) which recruited patients with severe COVID-19 with high oxygen requirement or mechanical ventilation from May to October 2020. The primary outcome, death at 28, was analyzed.

**RESULTS:** Between May and October 2020, 196 predominantly male patients (n=122, 62.2%) with an average of 58.1 years (± 15.5), were included in the cohort. Mortality at 28 days was 44.3 % (n= 84). Patients included in the second trimester had a greater mortality rate when compared with those recruited in the first trimester (54.1 *vs* 32.1, p< 0.01). On multivariate analysis, the detected protective factors were the use of fentanyl HR 0.51 (95%CI 0.31 – 0.85, p=0.01), the use of antibiotics HR 0.22 (95% CI 0.13 – 0.36, p<0.01), and a previously healthy state (no comorbidities other than obesity) HR 0.58 (95%CI 0.35 – 0.94, p =0.03); risk factors were severe kidney injury (AKIN3) HR 1.74 (95%CI 1.04 – 2.9, p=0.035), elevated D-Dimer levels HR 1.02 (95%CI 1.007 – 1.04, p=0.005), shock OR 5.8 (2.4 – 13.8, p<0.01), and recruitment in the second trimester OR 2.3 ((1.1 – 4.8, p=0.02).

**CONCLUSIONS:** In-hospital mortality in critically ill COVID-19 patients has increased in our center. The appropriate use of antibiotics, the type of sedation, and AKIN3 are modifiable factors directly related to this increased mortality. The increase in mortality observed in the second trimester is explained by hospital overcrowding that began in August 2020.

## Introduction

A little over a year ago, the first cases of COVID-19 were reported in China (1). Initial reports referred that 5 % of cases required invasive mechanical ventilation (IMV). (2) IMV-associated mortality is very high, reported as 56% in a recent meta-analysis of 69 studies (CI95% 47 – 65%) (3). In Mexico, IMV-associated mortality is 68%, based on official data (CI95% 67 - 70) (4).

Various non-modifiable risk factors have been associated with this high mortality (5). An interesting reported characteristic was a decrease in mortality in the initial phases of the pandemic, from 42% to 20 %, perhaps the result of a better-prepared health system (6,7). Mexico is the country with the highest lethality rate per 100 confirmed cases (8.7 %), and 109.2 per 100 thousand inhabitants. (8) However, this lethality rate occurs in a country in which only 17 tests are applied per 100 thousand inhabitants, among the lowest testing numbers in the region (9).

The Mexican health system is fragmented and is generally divided into public and private. In turn, the public system is further divided, depending on employment, into 5 sub-systems. An open population health system is another available alternative independent of employment. Each of these health systems for workers is financed differently, and its human resources are also different. This generates heterogeneous care scenarios that when faced with a disease as severe as COVID-19, directly compromise its prognosis.

We aim to evaluate the risk factors associated with mortality at 28 days in critically ill COVID-19 patients in a public hospital in Mexico.

## Methodology

This is a retrospective analysis of the patients included in the clinical trial (NCT04381858) which recruited patients with severe COVID-19 with high oxygen requirement or mechanical ventilation from May to October 2020. The methodology is detailed extensively in the original study. Briefly, we designed a controlled, randomized, open clinical trial that included patients with severe pneumonia secondary to SARS-CoV-2 infection, with severe respiratory failure or warranting IMV. Patients were randomized in a 2:1 ratio to receive either COVID-19 convalescent plasma, one 200 ml unit every 24 hours administered in two doses, or human immunoglobulin, 0.3 mg/kg/ day for 5 days. Patients were followed until hospital discharge or death.

We included patients fulfilling the following criteria: a) A positive SARS-CoV-2 RT-PCR test by nasal or oropharyngeal swab, b) Pneumonia diagnosed by X-ray or high-resolution chest CT scan, with a pattern suggesting coronavirus involvement, c) Severe hypoxemic respiratory failure of recent presentation or acute clinical exacerbation of pre-existing lung or heart disease requiring high oxygen levels (reservoir mask > 10 L/ min), high-flow nasal cannula (FiO2 > 80% 60 liters per minute) or IMV. Patients were excluded if pneumonia appeared not to be due to SARS-CoV-2, or if SARS-CoV-2 infection was not suspected to be the cause of respiratory deterioration.

On admission, we obtained blood samples to determine arterial blood gases, a complete blood count, blood chemistry, and prognostic markers such as fibrinogen, D-dimer, ferritin, troponin I, and C-reactive protein.

All hospitalized patients were administered thromboprophylaxis with low-molecular-weight heparin or unfractionated heparin, according to local and international guidelines (10). On the last week of June and based on the evidence obtained in the RECOVERY trial, we began to administer dexamethasone, 6 mg intravenously, every 24 hours for 10 days or until discharge (11).

Patients were also administered ivermectin, 12 mg, if their weight was below 80 kg, and 18 mg in case it was above 80 Kg; this treatment was established in all patients since, at the time, it appeared to have theoretical therapeutic potential on the basis of institutional guidelines (06/03/2020). A clinical trial was simultaneously conducted in non-critical patients requiring hospitalization, to compare the efficacy and safety of ivermectin and hydroxychloroquine (Clinical Trials Identifier: NCT04391127). In August, its analysis proved that both drugs were therapeutically futile, so ivermectin administration was suspended in the protocol patients (08/16/2020). (12)

Patients were discharged once they fulfilled the following criteria: absence of neurological complications, no fever, hemodynamic stability for at least 72 hours, minimal oxygen requirements (nasal prongs at 1-2 liters per minute), and a well-established support system.

This study was conducted at the *Hospital Centenario Miguel Hidalgo* in the state of Aguascalientes (Mexico), a tertiary care institution for the population lacking social security.

The original clinical trial was approved by the ethics committee of the *Hospital Centenario Miguel Hidalgo* on April 15 2020, with the registration number 2020-R-25. It was also uploaded in the ClinicalTrials.gov portal with the identifier NCT04381858.

## Statistical Analysis

Descriptive statistics were used according to the measurement level. The distribution of continuous variables was evaluated with the Kolmogorov Smirnov test. Continuous variables with a normal distribution were expressed as means and standard deviation, while those with an abnormal distribution were expressed as medians and interquartile intervals. Categorical variables were expressed as relative and absolute frequencies. In between-group analyses, according to the distribution of continuous variables, were established with Student’s t or Mann Whitney’s U. Dichotomic or ordinal variables were analyzed with the Chi^2^ test or Fisher’s exact test, as needed. Overall survival analysis for the outcome “death at 28 days” was conducted with Kaplan-Meier curves, and between-group comparisons were established with the Log-rank test. Multivariate analysis was performed with the Cox proportional hazards model and logistic regression. These analyses included variables with a p value ≤ 0.1, those with biological relevance, and the lack of a confounding phenomenon. A p value below 0.05 was considered significant. Analyses were conducted with Microsoft Excel 2013 and STATA version 11.1 software.

## Results

Between May 5, 2020 and October 17 of the same year, we included 196 patients with an average age of 58.1 years (± 15.5), most of whom were male (62.2 %). The most frequent comorbidity was a history of smoking (41 %), followed by obesity and overweight (40.8% and 38.7 %, respectively), systemic arterial hypertension (SAH) (35.7 %), and type 2 diabetes mellitus (DM2) in 34.7 % of cases. Other than the obesity and overweight comorbidities, 37.2 % (n=73) had no other pathologies, and only 6 patients had no comorbidities (Table 1).

**Table 1.**
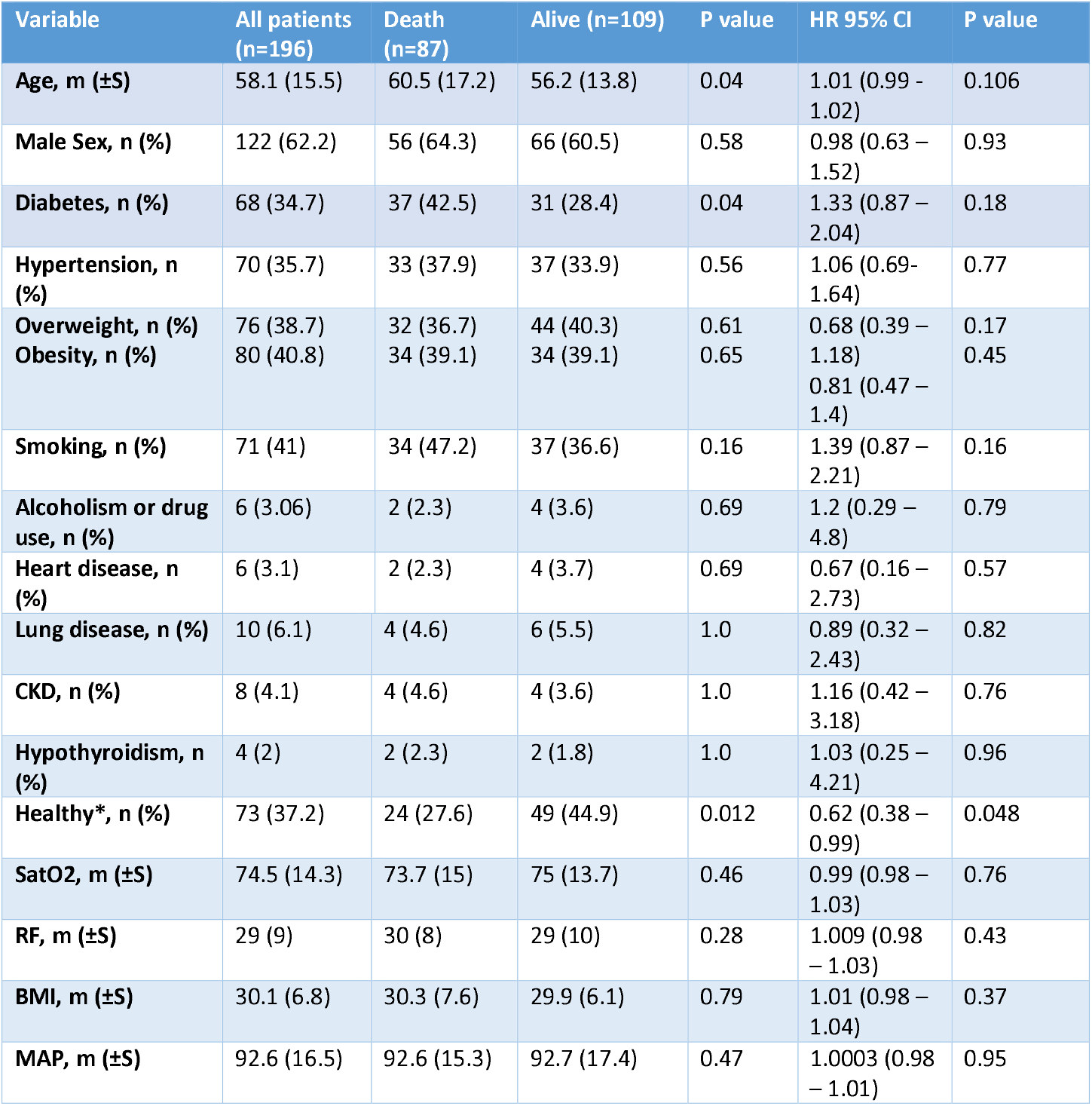
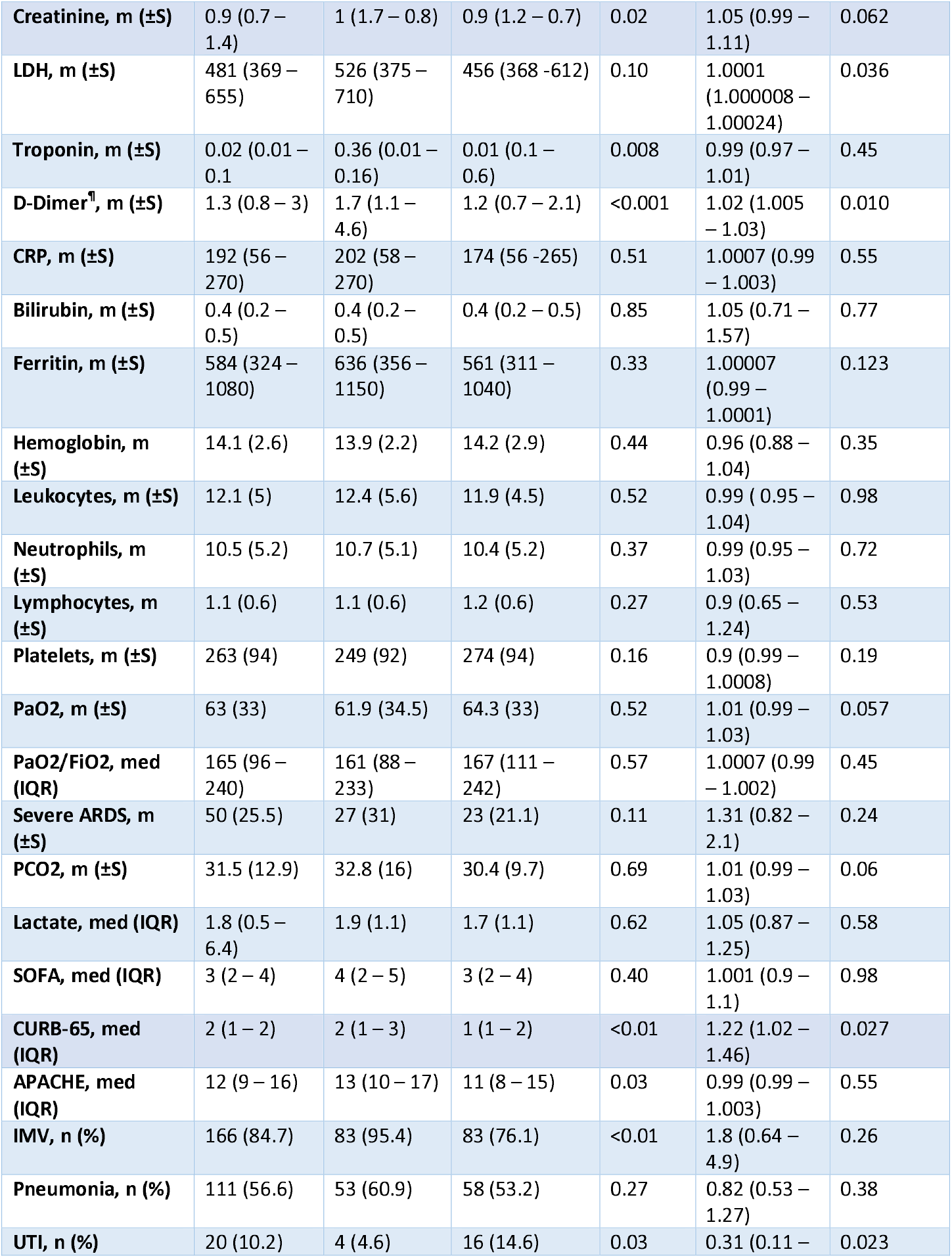
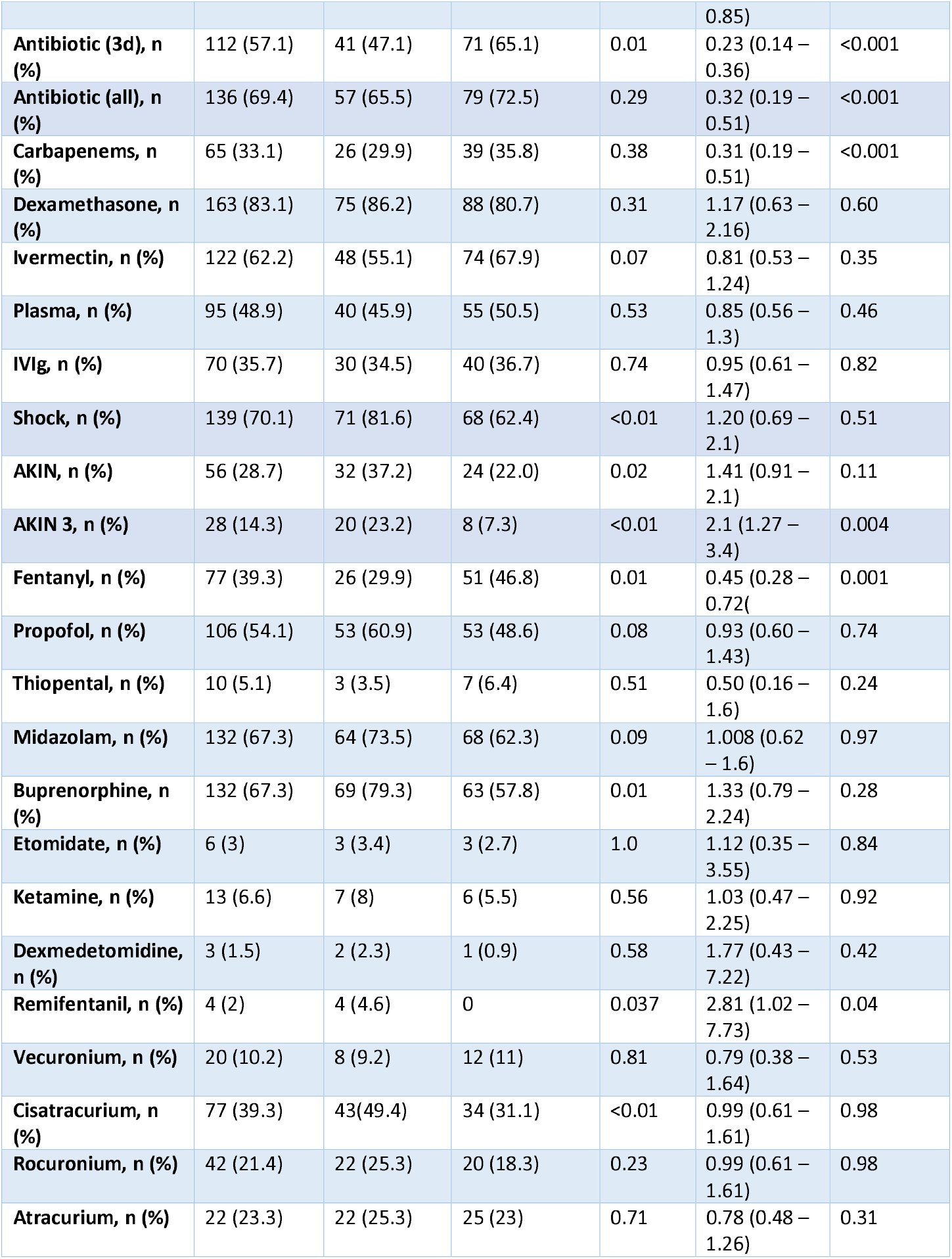
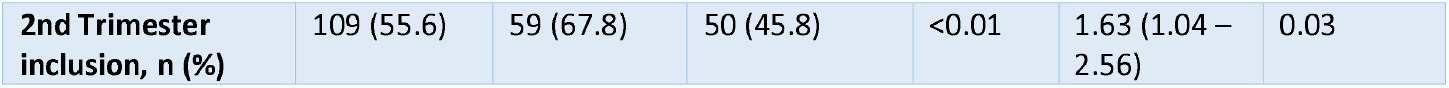
General characteristics, between-group mortality analysis, and Cox proportional hazards univariate analysis. *Healthy: Defined as the absence of comorbidities other than overweight and obesity ¶: D-Dimer expressed in thousands. IVIg: Intravenous Immunoglobulin, AKIN: Acute Kidney Injury, UTI: Urinary Tract Infection, CRP: C-reactive protein, LDH: Lactic dehydrogenase, RF: Respiratory Frequency, BMI: Body Mass Index, MAP: Mean arterial pressure, CKD: Chronic Kidney Disease.

At inclusion, 50 patients (25.5 %) had an oxygenation index under 100, 74 (37.7 %) between 100 and 200, and 51 (26 %) between 200 and 300. Average mean arterial pressure (MAP) at admission was 92.6 mmHg (± 16.5), and only 11 patients (5.6%) had a MAP below 65 mmHg.

Severity indices on admission included a median SOFA of 3 (2–4), CURB-65 2 (1 – 2), and APACHE II of 12 (9 – 16). IMV was warranted in 84.7 % (n= 166) of patients.

During follow-up, 70 % (n= 139) of patients developed a state of shock, 56.6 % (n=111) were diagnosed with mechanical ventilation-associated pneumonia, and 10.2 % (n=20) developed urinary tract infection (UTI). Antibiotic therapy was prescribed to 57 % (n=112) of patients for more than three days, and 69 % (n=136) received at least one dose of antibiotics. Some degree of acute kidney injury (AKIN) developed in 28.7 % (n= 56) of cases, and in 14.3 %, injury was severe (AKIN-3).

During the first 28 days of hospitalization, there were 87 deaths (44.3 %), among which only 8 were not the result of sepsis or adult respiratory distress syndrome. Four patients developed arrhythmias or acute myocardial infarction, two had severe pancreatitis and one had a stroke. The median time until death was 10 days (IQI 6 – 15).

## Mortality Analysis

Patients included during the first trimester of the study had a lower mortality compared with those enrolled in the second trimester (32.1% *vs* 54.1%, p< 0.01). This was also reflected in the survival analysis (Figure 1-A), and the difference was significant (p=0.02).

**Figure 1.**
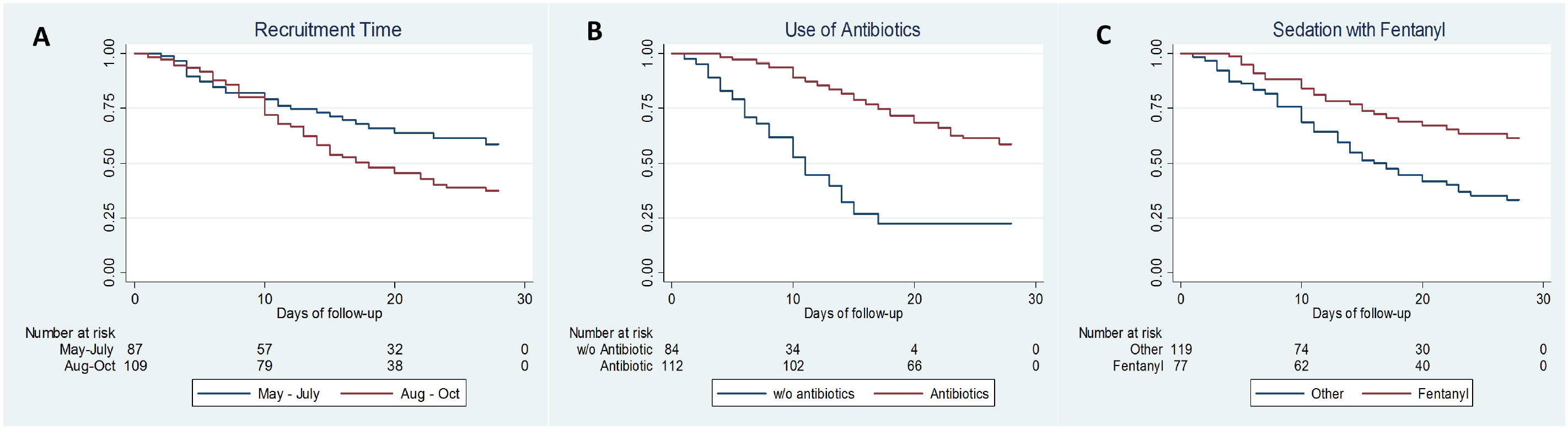
**1-A** Mortality at 28 days, estimation by Kaplan Meir according to the study inclusion trimester, Log-Rank p= 0.02. **1-B**. Survival Analysis of patients on antibiotic(s) (at least for 3 days) Log Rank p<0.01 **1-C**. Survival Analysis of patients on Fentanyl *vs* other sedation strategies, Log Rank p<0.01.

Mortality risk factors are described in Table 1 and Figure 1. Multivariate analysis with Cox proportional hazards adjusted to the inclusion trimester showed that protective factors were the lack of comorbidities other than overweight and obesity and the use of antibiotics and fentanyl as a sedative. Risk factors were acute kidney injury AKIN-3 and elevated D-dimer concentrations (Table 2).

**Table 2.** Multivariate analysis by Cox proportional hazards and logistic regression. *Adjusted to the inclusion trimester

| Cox Proportional Hazards* |  |  | Logistic Regression |  |
| --- | --- | --- | --- | --- |
| Variable | HR (95% CI) | P value | OR (95 % CI) | P value |
| **Fentanyl** | 0.51 (0.31 – 0.85) | 0.010 | 0.39 (0.17 – 0.85) | 0.018 |
| **Healthy** | 0.58 (0.35 – 0.94) | 0.030 | 0.47 (0.23 – 0.93) | 0.032 |
| **Use of Antibiotic** | 0.22 (0.13 – 0.36) | <0.001 | 0.29 (0.14 – 0.61) | 0.001 |
| **AKIN 3** | 1.74 (1.04 – 2.9) | 0.035 |  |  |
| **D-Dimer** | 1.02 (1.007 – 1.04) | 0.005 | 1.05 (1.002 – 1.1) | 0.039 |
| **Second trimester** |  |  | 2.3 (1.1 – 4.8) | 0.02 |
| **Shock** |  |  | 5.8 (2.4 – 13.8) | <0.001 |

By logistic regression, protective factors were the use of fentanyl, the lack of comorbidities aside from overweight and obesity, and the use of antibiotics. Risk factors were the development of shock, D-dimer levels, and the trimester of inclusion into the study (Table 2).

The inclusion trimester led to an interactive phenomenon particularly in terms of the survival analysis. We conducted a sub-group analysis (first and second trimester). Overall, patients included in the first trimester had a lower number of comorbidities and illness severity than those enrolled in the second trimester. (Appendix 1)

## Discussion

COVID-19 associated mortality in critically ill patients in our center is high and similar to that previously reported throughout the country (4,13). However, unlike observations in other centers, mortality has significantly increased over time, from 32% between May and July to 54% between August and October. This may be partly explained by the presence of more associated comorbidities and greater disease severity in patients included in the second trimester (Appendix 1). Also, hospital oversaturation played a fundamental role. In August, the number of hospital admissions exceeded the capacity of the intensive care unit, and new areas for critically ill patients had to be adapted (Figure 2). In other hospitals in the country, this overcrowding has even precluded their ability to provide IMV to a proportion of patients initially admitted without warranting this form of therapy (14,15).

**Figure 2.**
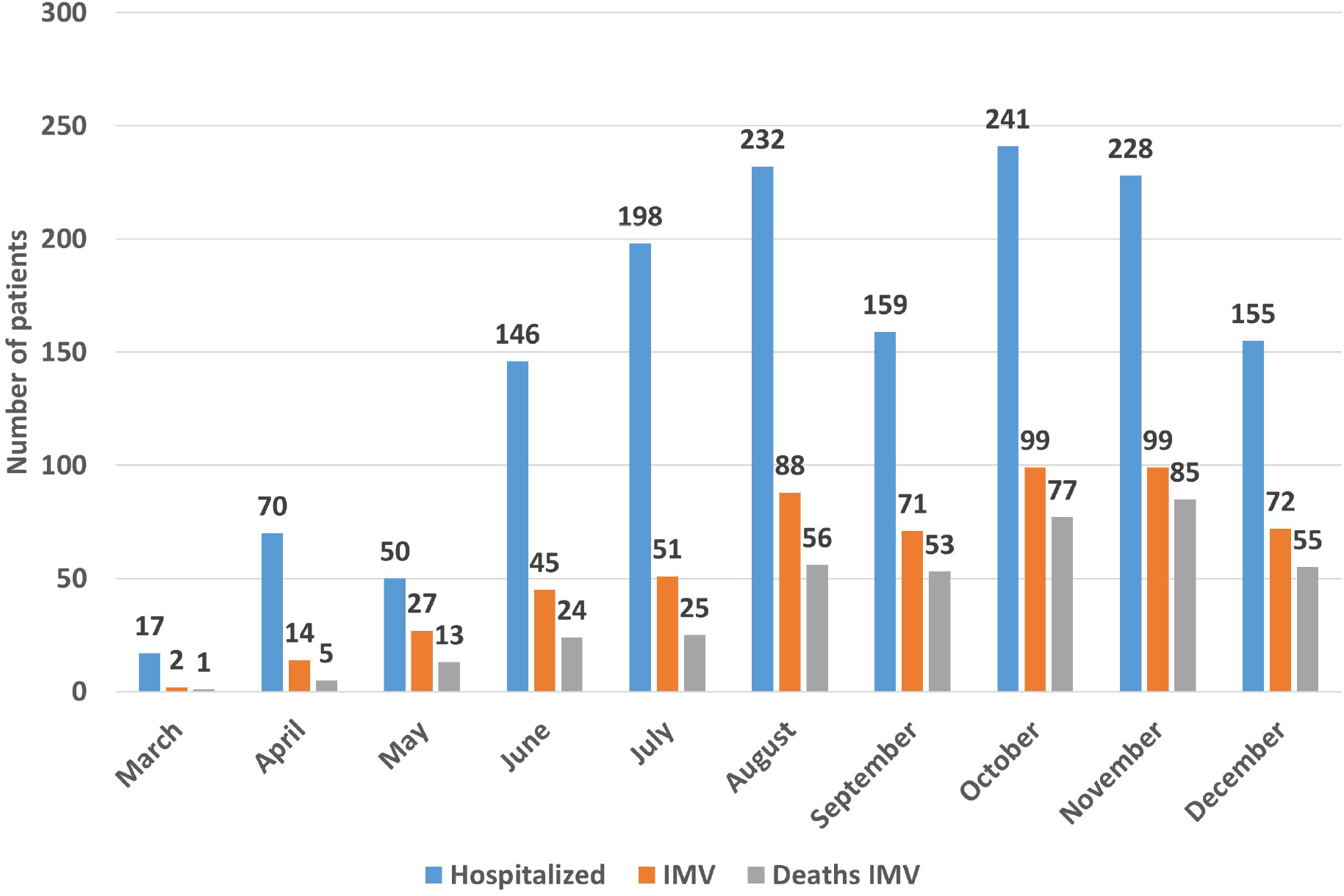
Number of hospitalized patients due to COVID-19 per month, those requiring IMV, and deaths on IMV.

The most relevant protective factor in terms of mortality was the use of antibiotics, which was independent of the trimester in which they were administered (56.8% *vs* 57.4 %, p =0.93). This is a concerning situation since the use of antibiotics is attributed to superinfections, a risk factor *per se*. Bacterial pneumonia developed in 56.6 % of patients (n=111), and 10.2 % (n=20) had a UTI. Fifty-seven percent (57%) (n = 111) of cases received antibiotics for at least 3 days, and 69.4 % (n =136) received at least one dose. This significantly differs from reports from other centers whereby a recent systematic review reported only 8% of bacterial coinfections, but most patients (72%) had received broad-spectrum antibiotics for unknown reasons (16). In an observational study in Mexico City, 98.5 % of cases were administered antibiotics without clearly establishing a rationale for their prescription (17). Appendix 2 describes the pattern of antibiotic use during the study more broadly.

A possible explanation for the improvement in mortality in our patients treated with antibiotics may hinge on local policies referring to the initiation of antimicrobials. Based on the lower proportion of bacterial coinfections reported in the literature, and prevent bacterial resistance, we vied to adopt a restrained conduct when beginning antibiotic therapy, attributing the inflammatory response and even the development of shock to viral activity; antibiotics were only administered if shock was refractory to treatment or cultures were positive (18). This probably delayed the timely initiation of antibiotics, one of the consistently related factors with death in a septic state (19).

Another mortality protective factor was the use of fentanyl, and this factor was directly related to the point at which the patient was included in the study; it was significantly greater in the first trimester. In the first months of the pandemic, appropriate drugs for sedation and analgesia were constantly scarce in our center. This paucity of drugs was not exclusive to our hospital or the country, since there is a general lack in the procurement and distribution of these drugs (20). The use of opioids for analgesia and patient sedation entails an adequate ventilatory status and shortens the duration of mechanical ventilation in conjunction with a safer hemodynamic profile (21–23). Several drug combinations were used in our center to obtain adequate sedation and analgesia but this form of management was never unified. In October, the use of thiopental became necessary although it is not recommended due to its safety profile (24). However, some centers do recommend it given the prolonged periods of intubation and the lack of more appropriate options (25).

Acute kidney injury (AKIN) has been solidly associated with mortality in intensive care patients (26,27). In hospitalized patients with COVID-19, the frequency of acute kidney injury is 10% and closely associated with mortality and severe infection (28–30). In our study, the prevalence of AKIN is very similar to that reported in critical patients in other centers (13,30). There are clinical practice guidelines and efforts that focus on the prevention of AKIN in critical patients, and that should be taken into account and evaluated in the current context (31,32).

COVID-19 continues to challenge our health system. Based on our center’s experience, the availability of ventilators and physicians trained in their use are not sufficient to guarantee superior patient outcomes. The reconversion of intensive care hospitalization areas in conjunction with a lack of epidemiology supervising personnel has led to a disturbing frequency of hospital-acquired infections. The scarcity of optimal sedative drugs has also compromised our ability to shorten the duration of mechanical ventilation and decrease the risk of infections.

Finally, the mortality described in this study is underestimated due to patients inclusion bias of clinical trials. In an analysis at our center on overall mortality in patients requiring IMV, data is much more alarming than that presented in this study (Figure 2).

## Conclusion

In-hospital mortality among critically ill COVID-19 patients has increased in our center. Based on the evaluated factors, we suggest the following:

a. Timely initiation of antimicrobial therapy following local epidemiology, in conjunction with contention measures and epidemiological surveillance to avoid the propagation of infections.
b. Search for alternatives to adequately sedate patients.
c. Increased vigilance of factors associated with the development of acute kidney injury.
d. Continue using potential treatments such as convalescent plasma or other therapies under investigation.
e. Increase the numbers and training of the personnel involved in patient management.

## Data Availability

All data is available upon request directly to the corresponding author

