## Appendix 1 for "Factors associated with increased mortality in critically ill COVID-19 patients in a Mexican public hospital: the other faces of health system oversaturation"

| MORTALITYCOMPARISON BETWEEN INCLUSION IN THE FIRST AND SECOND TRIMESTER Variable | First trimester  (n= 87) | Second trimester  (n= 109) | P Value |
| --- | --- | --- | --- |
| Age, med (IQI) | 55 (47 – 68) | 60 (47 – 74) | 0.21 |
| Sex, Male, n (%) | 62 (56.8) | 60 (68.9) | 0.08 |
| Diabetes, n (%) | 26 (29.8) | 42 (38.5) | 0.21 |
| SAH, n (%) | 60 (34.5) | 40 ( 36.7) | 0.74 |
| Stroke, n (%) | 2 (2.3) | 0 | 0.19 |
| Smoking, n (%) | 35 (43.7) | 36 (38.7) | 0.50 |
| Heart disease, n (%) | 3 (3.4) | 3 (2.7) | 1.0 |
| Lung disease, n (%) | 6 (6.9) | 4 (3.6) | 0.34 |
| Cancer, n (%) | 1 (1.1) | 0 | 0.44 |
| Transplant, n (%) | 1 (1.1) | 0 | 0.44 |
| Autoimmunity, n (%) | 0 | 2 (1.8) | 0.50 |
| Overweight, n (%) | 38 (43.6) | 38 (34.8) | 0.21 |
| Obesity, n (%) | 28 (32.1) | 52 (47.7) | 0.02 |
| Healthy, n (%) | 33 (37.9) | 40 (36.7) | 0.85 |
| MAP, m (S) | 93.9 (15.1) | 91.1 (18.1) | 0.25 |
| Creatinine, med (IQI) | 0.9 (0.7 – 1.4) | 0.9 (0.7 – 1.5) | 0.89 |
| LDH, med (IQI) | 479 (375 – 649) | 482 (367 – 658) | 0.91 |
| Troponin, med (IQI) | 0.02 (0.01 – 0.09) | 0.02 (0.01 – 0.11) | 0.34 |
| D-Dimer,med (IQI) | 1.4 (0.9 – 3.3) | 1.3 (0.7 – 2.7) | 0.63 |
| Ferritin, med (IQI) | 561 (324 – 1080) | 625 (333 – 1060) | 0.69 |
| SOFA, med (IQI) | 3 (2 – 4) | 3 (2 – 5) | 0.58 |
| APACHE, med (IQI) | 11 (9 – 14) | 13 (9 – 18) | 0.03 |
| CURB 65, med (IQI) | 1 (0 – 2) | 2 (1 – 3) | <0.01 |
| IMV, n (%) | 70 (80.4) | 96 (88.1) | 0.14 |
| T of IMV, med (IQI) | 13 (6 – 24) | 12 (7 – 23) | 0.76 |
| Fentanyl, n (%) | 51 (58.6) | 26 (23.8) | <0.01 |
| Ivermectin, n (%) | 76 (87.4) | 46 (42.2) | <0.01 |
| Shock, n (%) | 64 (73.5) | 75 (68.8) | 0.46 |
| AKIN (all), n (%) | 21 (24.1) | 35 (32.4) | 0.21 |
| AKIN 3, n (%) | 8 (9.2) | 20 (18.5) | 0.06 |
| Antibiotics (all), n (%) | 57 (65.5) | 79 (72.4) | 0.29 |
| Antibiotics 3d, n (%) | 50 (57.4) | 62 (56.8) | 0.93 |

**Supplementary Table 1:** Patient characteristics divided by trimesters (May – July *vs* August – September).
