## Appendix 2 for "Factors associated with increased mortality in critically ill COVID-19 patients in a Mexican public hospital: the other faces of health system oversaturation"

**Use of antibiotics**

A total of 112 (57 %) patients received antibiotics during follow-up. The average time to initiation of the first protocol lasting at least 3 days, was 7 days (IQI 2 – 11). The most frequently used antibiotics were carbapenems in 65 patients, in conjunction with levofloxacin in 18 cases. The period in which the first antibiotic protocol was administered was 6 days (IQI 4 – 8).

Patients on antibiotics were younger (54 *vs* 64 years, p <0.01), they had a greater number of total neutrophils (10.3 *vs* 9.6, p=0.03), a greater proportion developed shock (80.9 *vs* 58.1, p <0.01), and their hospitalization was more prolonged (22.5 *vs* 8 days, p <0.01) (Supplementary table 2).

Regardless of the week of their administration, patients who received antibiotics had a lower mortality rate than cases that did not receive antibiotics (Supplementary table 3).

**CULTURES**

In 196 patients, the average number of positive cultures was 2.5, with a median of 1 (IQI 0 to 4). Eighty-seven patients (44.6 %) had a positive bronchial secretion culture (min-max 1 – 11). There were 87 sputum isolates, with a predominance of Pseudomonas (n=44), Klebsiella Pneumoniae (n=32), S. aureus (n=14), and Acinetobacter (n= 13). Forty-three point 2 (43.2 %) of Pseudomona strains were carbapenem-resistant.

| Variable | Use of antibiotic  (n= 110) | No Antibiotic  (n= 86) | p Value |
| --- | --- | --- | --- |
| Sex Male, n (%) | 71 (64.5) | 51 (59.3) | 0.45 |
| Age, med (IQI) | 54 (47 – 67) | 64 (47 – 75) | <0.01 |
| Comorbidities  Diabetes Mellitus  SAH  Overweight  Obesity  Smoking  Heart disease  Lung disease  CKD  Healthy | 42 (38.2)  39 (35.4)  46 (41.8)  42 (38.1)  3 (2.7)  4 (3.6)  6 (5.4)  6 (5.4)  38 (34.5) | 26 (30.2)  31 (36)  30 (34.8)  38 (44.2)  4 (4.6)  2 (2.3)  4 (4.6)  2 (2.3)  35 (40.7) | 0.24  0.93  0.32  0.39  0.70  0.69  1.0  0.47  0.45 |
| PaO2/FiO2, med (IQI) | 160 (100 – 223) | 177 (95 – 247) | 0.51 |
| BMI, med (IQI) | 28 (25 – 32) | 28 (25 – 35) | 0.39 |
| MAP, med (IQI) | 90 (82 – 98) | 94 (83 – 104) | 0.08 |
| Creatinine, med (IQI)  LDH, med (IQI)  Troponins, med (IQI)  D-dimer, med (IQI)  Neutros, med (IQI)  Lymphos, med (IQI)  Lactate, med (IQI) | 0.9 (0.7 – 1.3)  499 (371 – 681)  0.029 (0.01 – 0.1)  1.2 (0.8 – 2.9)  10.3 (1.2 – 14.7)  1.04 (0.6 – 1.5)  1.6 (1.1 – 2.2) | 0.9 (0.7 – 1.5)  460 (344 – 590)   0.01 (0.01 – 0.1)   1.6 (0.8 – 3.2)  9.6 (6.2 – 11.7)  1.01 (0.71 – 1.57)  1.5 (1.1 – 1.8) | 0.81  0.13  0.08  0.69  0.03  0.86  0.41 |
| SOFA, med (IQI)  APACHE, med (IQI)  CURB65, med (IQI) | 3 (3 – 4)  12 (9 – 16)  2 (1 – 2) | 3 (2 – 4)  12 (9 – 16)  2 (1 – 3) | 0.21  0.75  0.51 |
| Dexameth, n (%)  Ivermectin, n (%)  Anticoagulation, n(%)  IVIg, n (%)  Plasma, n (%) | 92 (83.6)  65 (59)  109 (99)  41 (37.2)  55 (50) | 71 (82.5)  57 (66)  84 (97.6)  29 (33.7)}  40 (46.5) | 0.84  0.30  0.58  0.60  0.66 |
| Shock, n (%) | 89 (80.9) | 50 (58.1) | <0.01 |
| AKIN, n (%)  AKIN 3, n (%) | 35 (31.8)  15 (13.6) | 21 (24.7)  13 (15.3) | 0.27  0.83 |
| Outcomes  Death, n (%)  Hospitaliza, m (IIC)  Death at 28 d, n (%) | 55 (49)  22.5 (15 – 32)  41 (37.2) | 48 (57.1)  8 (15 – 32)  46 (53.4) | 0.26  <0.01  0.02 |

**Supplementary table 2.** General characteristics of patients on antibiotics for at least 3 days, those on antibiotics for less than 3 days, and those on no antibiotics.

| Scenarios | HR (95% IQI) | P value |
| --- | --- | --- |
| No Antibiotic | Ref | Ref |
| Antibiotic, first week | 0.39 (0.16 – 0.82) | 0.015 |
| Antibiotic, second week | 0.34 (0.18 – 0.64) | 0.001 |
| Antibiotic, third week | 0.36 (0.19 – 0.66) | 0.001 |
| Antibiotic after third week | 0.09 (0.04 – 0.22) | <0.01 |

*Adjusted to age and sex

**Supplementary table 3.** Groups on antibiotics according to the week the protocol was initiated in comparison with patients on no antibiotics.
